# Clioquinol improves catalytic activity of PHGDH and shows antiseizure efficacy in patients

**DOI:** 10.1101/2024.09.28.24314470

**Authors:** Karin Thevissen, Annelii Ny, Daniëlle Copmans, Jana Tits, Kenichi Kamata, Eva Gielis, Kevin Longin, Jo Sourbron, Peravina Thergarajan, Tracie Huey-Lin Tan, Idrish Ali, Nigel C. Jones, Terence J. O’Brien, Mastura Monif, Bridgette D. Semple, Christine Germeys, Benedetta Frizzi, Virginia Minniti, Sebastian Perrone, Carole L. Linster, Ilaria Elia, Ludo Van Den Bosch, Bruno P.A. Cammue, Arnout Voet, Lieven Lagae, Peter de Witte

**Affiliations:** Centre of Microbial and Plant Genetics, KU Leuven; Leuven, Belgium; Department Microbiology, Immunology and Transplantation, KU Leuven; Leuven, Belgium; Laboratory for Molecular Biodiscovery, KU Leuven; Leuven, Belgium; Laboratory for Biomolecular modelling and Design, KU Leuven; Leuven, Belgium; Department Development and Regeneration, Section Pediatric Neurology, University Hospitals Leuven; Leuven, Belgium and Member of European Reference Network Epicare; Center for Medical Genetics, Ghent University Hospital, Ghent, Belgium; Department of Neuroscience, Monash University; Melbourne, Australia; Department of Medicine (Royal Melbourne Hospital), The University of Melbourne; Parkville, Australia; Department of Neurology, Alfred Health; Prahran, Australia; Department of Neurology, Royal Melbourne Hospital; Victoria, Australia; Department of Neurosciences, and Leuven Brain Institute; KU Leuven, Leuven, Belgium; VIB, Center for Brain & Disease Research; Leuven, Belgium; Department of Cellular and Molecular Medicine; KU Leuven, Belgium; Luxembourg Centre for Systems Biomedicine, University of Luxembourg; Belvaux, Luxembourg

## Abstract

Drug-resistant epilepsy (DRE) affects over 25 million people worldwide and is associated with neuroinflammation. We identified haloquinolines from a drug repurposing library as potent activators of phosphoglycerate dehydrogenase (PHGDH) enzyme, which converts 3-phosphoglycerate to generate serine and the neurotransmitter glycine, and steers anti-inflammatory responses. The most promising haloquinoline clioquinol can increase the catalytic activity of PHGDH up to 2.5-fold, thereby increasing de novo glycine biosynthesis and resulting in reduced glutamate levels. Moreover, we show that clioquinol has PHGDH-dependent antiseizure activity as well as anti-inflammatory properties *in vivo* using various zebrafish and mouse epilepsy models. Finally, we demonstrate the efficacy of clioquinol as add-on treatment in severe DRE patients. Therefore, increasing activity of PHGDH is a promising new approach to treat DRE.

## Main Text

Epilepsy is a prevalent neurological disorder affecting over 80 million individuals worldwide (*1*), often accompanied by neuroinflammation that can both cause and result from epileptic activity (*2*). Antiseizure medications (ASMs) are commonly used to control seizures, while anti-inflammatory treatment is used in certain epilepsy syndromes, such as infantile spasms (*3*). However, approximately one third of patients are classified as having drug-resistant epilepsy (DRE), as they do not adequately respond to current ASMs despite the availability of more than 25 drugs on the market (*4*).

In this study, we investigated the enzyme phosphoglycerate dehydrogenase (PHGDH), mainly expressed in astrocytes in the brain (*5*), as a novel therapeutic target for treating DRE. By releasing so-called gliotransmitters such as glutamate upon an increase in intracellular Ca^2+^ into the tripartite synapse, astrocytes directly interfere with neuronal activity. Mounting evidence attributes an important role to astrocytes in the initiation and progression of epilepsy and concomitant neuro-inflammatory processes. However, targeting gliotransmission as an antiseizure treatment has not been reported (*6*). PHGDH catalyzes the conversion of the glycolytic intermediate 3-phosphoglycerate to 3-phosphonooxypyruvate (3-PHP), which is the rate-limiting step in *de novo* serine/glycine biosynthesis (Fig. 1A) (*7*). Subsequently, 3-PHP is converted into O-phospho-L-serine by phosphoserine aminotransferase 1 (PSAT1) in a glutamate-linked transamination reaction, and finally into L-serine by phosphoserine phosphatase (PSP). Ultimately, hydroxymethyltransferase (SHMT) converts L-serine into L-glycine. Increasing evidence suggests a link between the induction of seizures and (i) PHGDH dysfunction as a result of genetic mutations in the PHGDH gene (*8*, *9*) or (ii) PHGDH inhibition (*10*, *11*). Additionally, PHGDH has been identified as a critical enzyme in driving macrophage polarization towards an anti-inflammatory state (*12*). As a result, we hypothesized that PHGDH activators may be beneficial for treating DRE by exhibiting both antiseizure and anti-inflammatory activity. The metabolite 2-phosphoglutarate was previously shown to activate PHGDH (*13*), indicating that the concept of screening for novel PHGDH activator drugs is feasible.

**Fig. 1.**
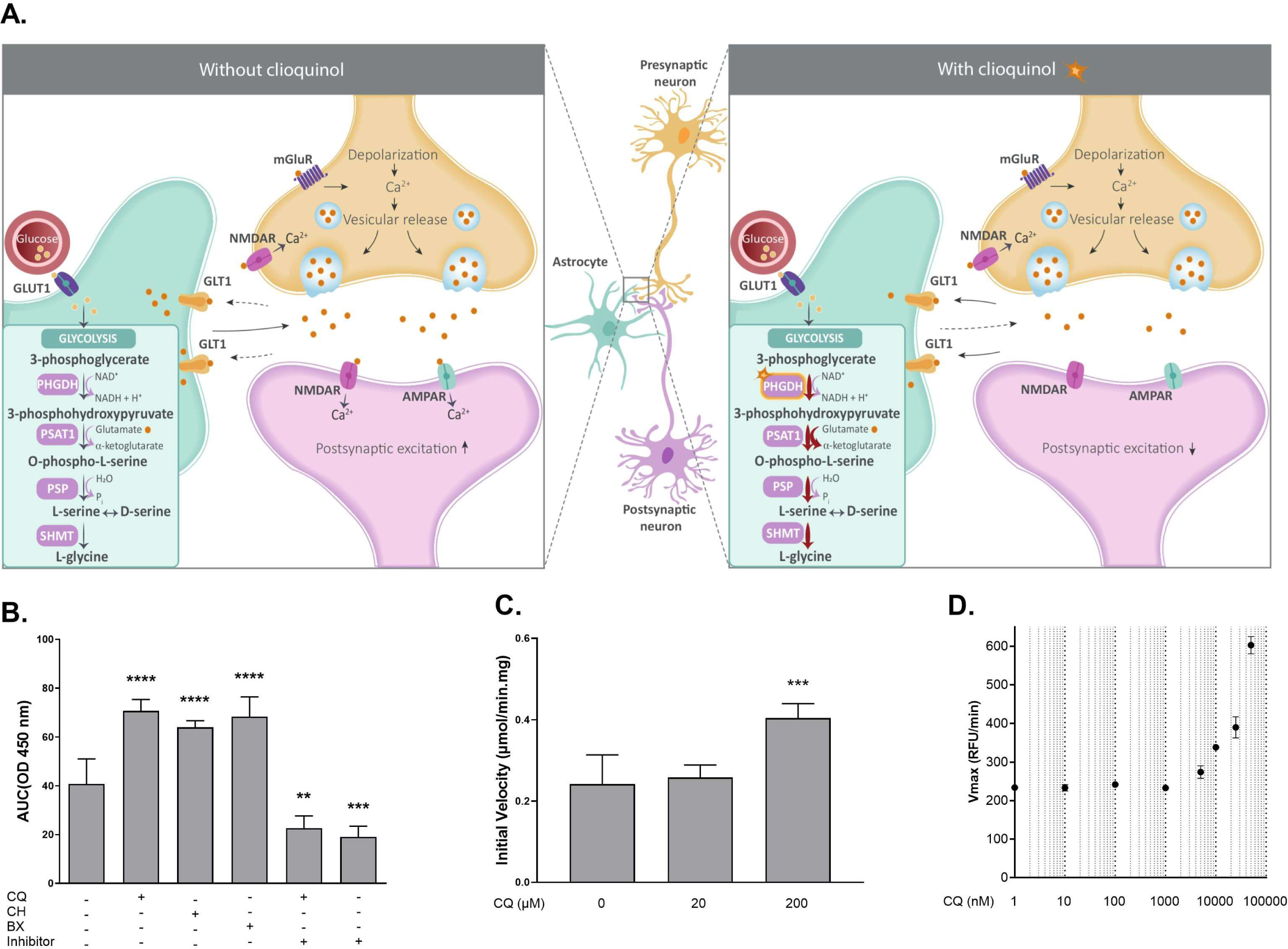
PHGDH activation by haloquinolines. (**A**) Schematic representation of *de novo* glycine/serine biosynthesis. Clioquinol activation of PHGDH results in increased 3-PHP and downstream glycine synthesis, involving PSAT1’s conversion of glutamate to α-ketoglutarate. (**B**) PHGDH activity with or without 25 µM chloroxine (CH), 25 µM broxyquinoline (BX) or 25 µM clioquinol (CQ), alone and for CQ also in combination with 10 µM of the PHGDH inhibitor CBR-5884. PHGDH activity was measured during 1h with OD450 nm as readout in a colorimetric assay, representing NADH generated. Data are shown as mean area under the curve (AUC) ± SD in at least 3 biological repeats. (**C**) NADH generated by PHGDH upon incubation with 20-200 µM CQ in the presence of deproteinized cell extract was measured spectrophotometrically. Data shown are means ± SD (n ≥ 5 in 2 biological repeats). Statistical differences: ****p<0.0001, ***p<0.001, **p<0.01 and *p<0.05 by one-way ANOVA with Dunnett’s multiple comparisons test (**B, C**). (**D**) Vmax (kcat) of sPHGDH in the presence of different concentrations of CQ. Vmax was calculated based on colorimetric change due to NADH-induced resazurin to resorufin conversion at increasing (0.5-5 mM) 3-PG and 100-fold excess NAD^+^. Data are means ± SD of 3 biological repeats.

Here we identified several haloquinolines as PHGDH activators, with clioquinol (CQ), an antifungal and antiprotozoal drug (*14*), showing the most promising results. We assessed the antiseizure activity of CQ in zebrafish epilepsy models and the mouse 6-Hz psychomotor seizure model, and its anti-inflammatory properties in the self-sustained status epilepticus (SSSE) mouse epilepsy model. Additionally, the mechanistic consequences of activating PHGDH in a PHGDH-dependent cell line and in human astrocytes as well as in zebrafish heads were explored. Finally, we assessed the efficacy of CQ as add-on treatment in severe DRE patients in a clinical open pilot proof-of-concept study.

### Haloquinoline clioquinol increase catalytic activity of human PHGDH

A novel function of PHGDH in reducing reactive oxygen species (ROS) levels via increased glutathione synthesis was uncovered in 2020 (*15*). We previously identified various pharmaceutical compounds from a drug repurposing library that increased survival of yeast under lethal ROS-inducing stress (*16*). Tanshinone was one of the hits from this screening and is known to increase the expression of *Phgdh* (*17*), which is consistent with a role for PHGDH in the normalization of ROS levels. Hence, we assessed the potential activation of human PHGDH by all previously identified hits from the screening (*16*) and found that the haloquinolines clioquinol, chloroxine, and broxyquinoline resulted in increased PHGDH activity in an enzyme assay using human recombinant PHGDH and a NADH-based colorimetric readout (Fig. 1B). Clioquinol (CQ) was the most promising hit for further analysis, as it has been used clinically for a long time (*14*). Co-administration of the PHGDH-specific inhibitor CBR-5884 (*18*) with CQ completely blocked the induced PHGDH activation (Fig. 1B), indicating the PHGDH specificity of CQ’s readout in the assay. Next, we assessed the effect of CQ on human recombinant PHGDH in an enzymatic assay using conditions that mimic a cellular context, i.e. in the presence of PSAT1, the enzyme downstream of PHGDH, and a deproteinized cellular extract obtained from a HAP1 cell culture. The reaction was followed by measuring NADH spectrophotometrically and showed increased PHGDH activity by CQ in these conditions as well (Fig. 1C).

Previous kinetic studies with human PHGDH indicated that a truncated PHGDH variant, obtained by removal of the 2 regulatory C-terminal domains, yielded an enzyme (sPHGDH) with similar catalytic efficiency as full length protein (*19*). We performed further enzymatic studies with sPHGDH to limit the impact of allosteric regulation, and in the presence of EDTA to exclude effects due to metal-chelation by CQ (*20*). Activation of sPHGDH by CQ was observed from the turnover of 3-PG and NAD^+^ at CQ concentrations up to 50 µM (fig. S1). Further kinetic studies performed with varying 3-PG concentration and surplus NAD^+^ demonstrated a concentration-dependent increase of the maximal reaction rate V_max_ (from 239.4 ± 12.7 RFU/min to 611.7 ± 22.6 RFU/min; Fig. 1D), implying a 2.5-fold improvement of the catalytic constant K_cat_. As no plateau of V_max_ could be reached due to low solubility of CQ, the EC50 of CQ could not be determined. These results prove that CQ directly engages with PHGDH via the active site domains of PHGDH and increases its activity.

### Clioquinol enhances de novo glycine biosynthesis and results in decreased glutamate levels

By increasing the activity of PHGDH, we hypothesize that more 3-PHP is produced, which in turn will be converted to more o-phospho-L-serine by PSAT1 and ultimately, to more serine and/or glycine. As PSAT1 utilizes glutamate (*21*), which is an excitatory neurotransmitter (*22*), PHGDH activation should also result in reduced levels of intracellular glutamate. Hence, we measured intracellular levels of serine, glycine and glutamate and performed 13C6 glucose tracing in CQ-treated and untreated 4T1 breast cancer cells, which have a high dependency on serine/glycine biosynthesis, in serine- and glycine-free medium. CQ treatment of 4T1 cells combined with 13C6 glucose tracing revealed no difference in intracellular serine levels and glucose contribution to serine (Serine (M3)) compared to 1% DMSO control, excluding an increase of serine biosynthesis levels upon CQ treatment (Fig 2A). However, the same treatment resulted in an increase of intracellular glycine levels, significant at 7.5 µM CQ, coupled with a higher glucose contribution to glycine (Glycine (M2)), significant at both 5 µM and 7.5 µM CQ. These findings suggest an increased glycine biosynthesis upon CQ treatment. Additionally, the significant decrease in glutamate intracellular levels observed in the 7.5 µM CQ condition confirms the increased usage of this metabolite by PSAT1 to ultimately support glycine metabolism.

**Fig. 2.**
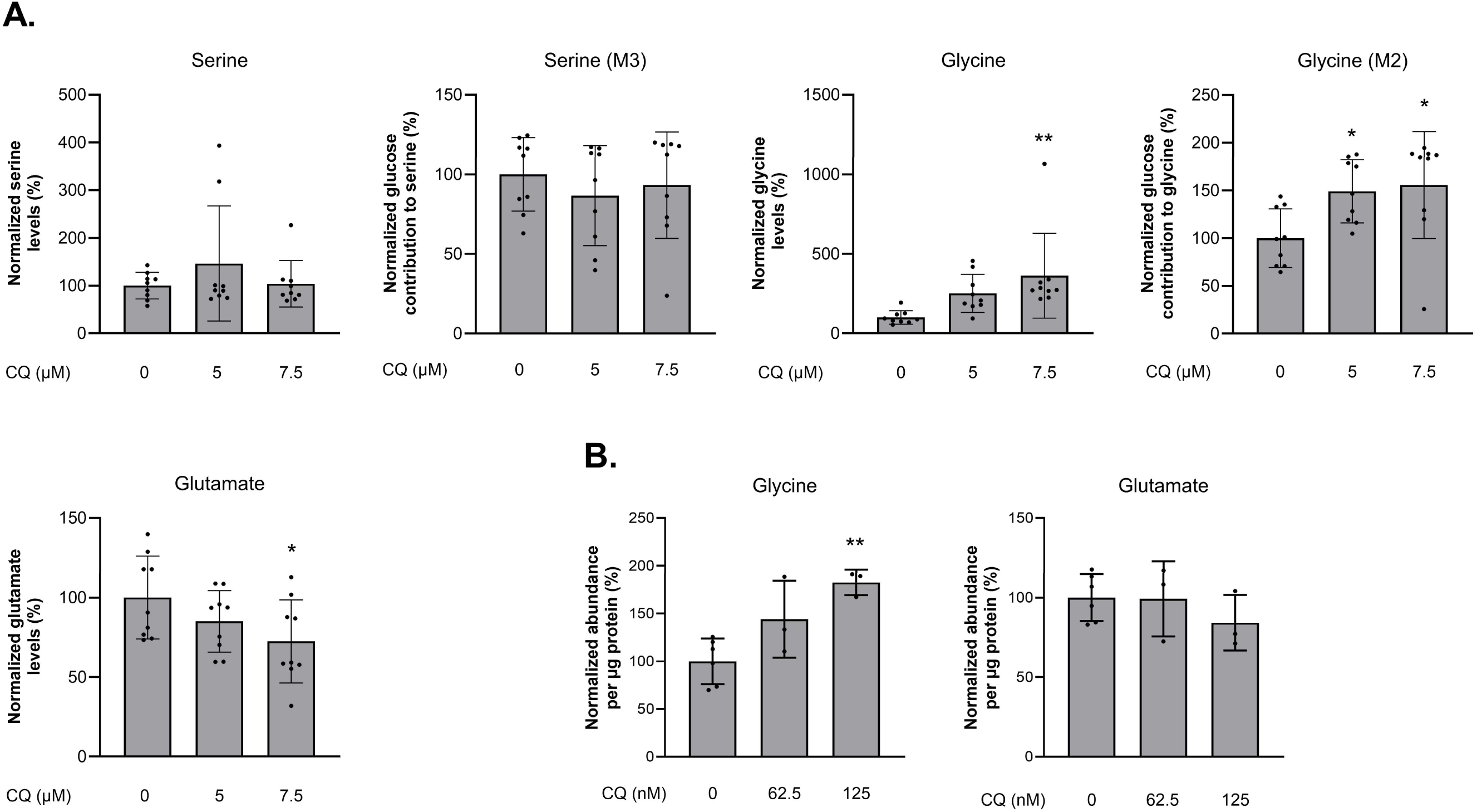
Impact of Clioquinol on the PHGDH-driven de novo glycine biosynthesis on 4T1 breast cancer cells and iPSC-derived astrocytes. **(A)** 4T1 breast cancer cells were incubated with 0 µM (n=9), 5µM (n=9) and 7.5µM CQ (n=9) in serine- and glycine-free medium in 3 biological repeats. Intracellular levels of serine, glycine and glutamate were measured using Gas Chromatography-Mass Spectrometry and the fraction of glucose in serine and glycine upon 13C6 glucose tracing was calculated and normalized to vehicle (0 µM) ± SD. (**B**) Intracellular glycine and glutamate abundances per µg protein ± SD in iPSC-derived astrocytes treated with vehicle (0 nM, n=6), 62.5 nM (n=3) or 125 nM CQ (n=3) for 50 h in 3 separate astrocyte differentiations. Statistical differences: **p<0.01 and *p<0.05 by one-way ANOVA with Dunnett’s multiple comparisons test.

Next, as PHGDH is mainly expressed in astrocytes in the brain, we subsequently assessed glycine and glutamate levels in human induced pluripotent stem cell (iPSC)-derived astrocytes treated with CQ. In line with the breast cancer cell line data, we found that treatment of astrocytes with CQ resulted in significantly increased intracellular glycine levels compared to the vehicle treated controls, as well as in a trend toward reduced levels of intracellular glutamate (Fig 2B).

Overall, these data indicate that CQ can affect the levels of two key neurotransmitters, glycine and glutamate, through the activation of PHGDH. Glycine is a major inhibitory neurotransmitter in the central nervous system, alongside gamma-aminobutyric acid (GABA) (*23*). Significantly, activation of glycine receptors has been shown to exert an anticonvulsive effect in mature rat hippocampus (*24*). Glutamate plays a critical role in excitatory neurotransmission resulting in cellular and network hyperactivity. Notably, it has been shown that, when dysregulated, glutamatergic neurotransmission is fundamental to epileptogenesis and seizures (*25*). Hence, a compound that can increase intracellular levels of glycine and decrease levels of glutamate bears great potential as an ASM.

### Clioquinol shows antiseizure activity in chemical and genetic zebrafish seizure models

Next, we used the pharmacologically validated zebrafish ethyl ketopentenoate (EKP) chemical seizure model (*26*) and the Dravet syndrome genetic epilepsy model (*27*) to investigate the activity of CQ against drug-resistant seizures. First, we confirmed the reported link between PHGDH dysfunction and seizures in zebrafish, using the PHGDH-specific inhibitor CBR-5884. We assessed the effect of CBR-5884 at its maximum tolerated concentration (MTC; i.e. 1 µM) on 7 days post fertilization (dpf) zebrafish larvae, on epileptiform brain activity via local field potential (LFP) recording (fig. S2). In line, we observed a significant increase in epileptiform brain activity in zebrafish wild-type larvae treated with CBR-5884.

Next, we assessed the effect of CQ (MTC of 1 µM) on locomotor and epileptiform brain activity in the zebrafish EKP seizure model. CQ significantly reduced EKP-induced locomotor activity (Fig. 3A) as well as EKP-induced epileptiform brain activity (Fig. 3B, fig. S3). Neither the locomotor activity nor the epileptiform brain activity of zebrafish larvae treated with CQ alone (without EKP addition) differed from vehicle (VHC)-only treated larvae. In contrast to 0.5 µM CQ alone, co-administration of CQ and CBR-5884 at the MTC for this combination (i.e. 0.64 µM CBR-5884) did not modify epileptiform activity induced by EKP (Fig. 3B), suggesting a PHGDH-dependent antiseizure action of CQ. The level of epileptiform activity upon treatment with 0.64 µM CBR-5884, followed by the addition of EKP, did not differ significantly from that of larvae treated with EKP alone. None of the compounds or combinations resulted in a significant increase or decrease in epileptiform activity in the absence of EKP compared to the VHC control treatment.

**Fig. 3.**
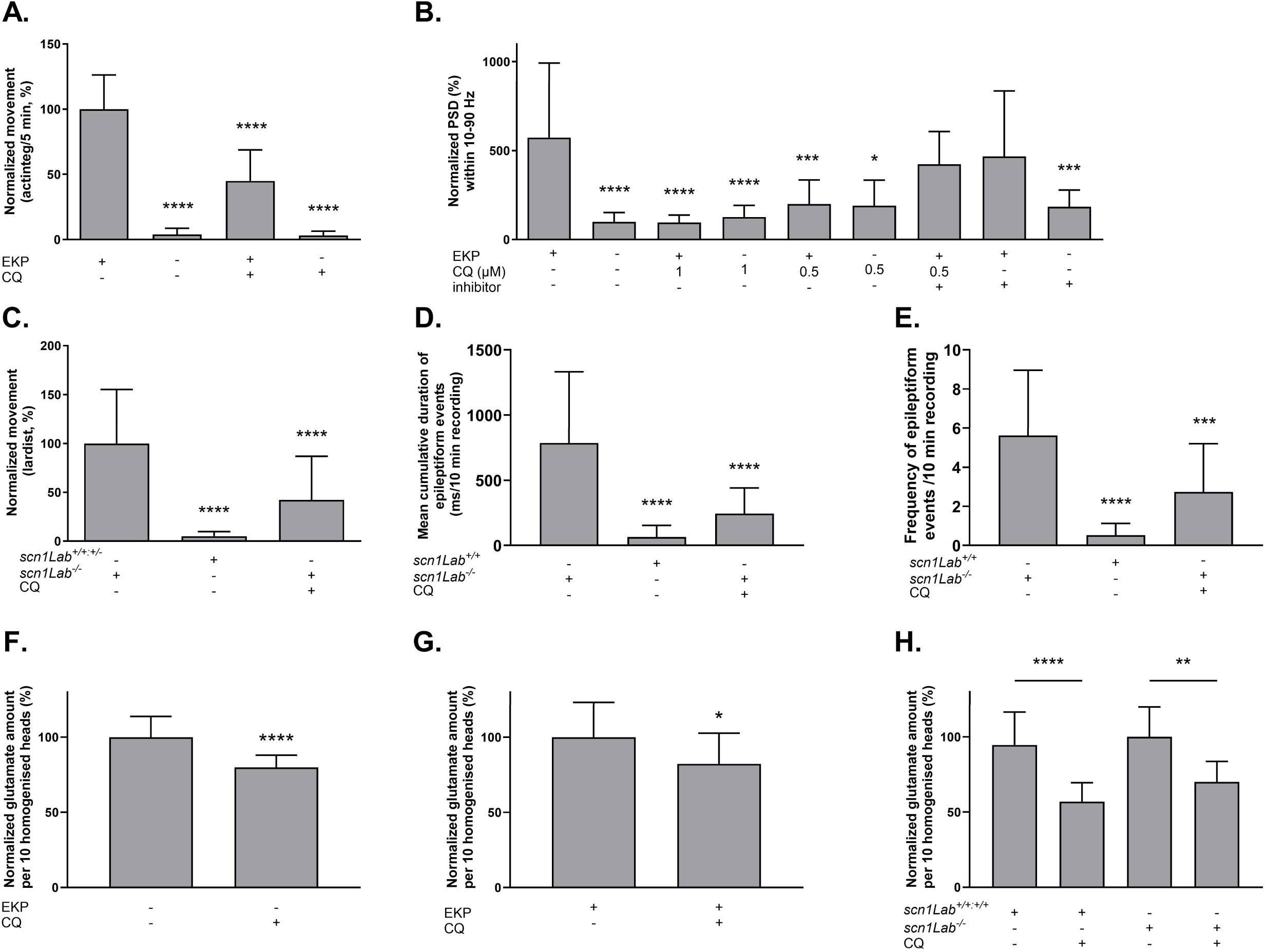
Antiseizure activity of clioquinol (CQ) in EKP and Dravet syndrome (*scn1Lab^-/^*^-^) zebrafish epilepsy models. Locomotor activity of larvae in (**A**) response to EKP and (**C**) Dravet syndrome; with or without 1 µM CQ. Activity expressed as (**A**) mean actinteg units per 5-min ± SD during 30 min, relative to EKP-only treatment; or (**C**) normalized lardist per 100-sec ± SD, relative to VHC (1% DMSO). Electrophysiological activity of larvae expressed as (**B**) mean normalized power spectral density (PSD) within a 10-90 Hz frequency range per larva ± SD, relative to VHC; (**D**) mean cumulative duration of epileptiform events (ms/10 min recording) and (**E**) frequency of epileptiform events/10 min recording ± SD, relative to *scn1Lab^-/-^*. For each experiment >10 larvae were used in at least 3 biological repeats. Glutamate levels in heads of (**F**) wild-type and (**G**) EKP treated larvae, and in (**H**) Dravet syndrome zebrafish, with or without 1 µM CQ. Results expressed as normalized glutamate amount per 10 homogenised heads ± SD. Statistical differences: ****p<0.0001, ***p<0.001, **p<0.01 and *p<0.05 by one-way ANOVA with Dunnett’s multiple comparisons test (**A-E**), by unpaired Student’s t-test (**F, G**) and by two-way ANOVA with Šídák’s multiple comparison test (**H**).

We further assessed the effect of CQ in Dravet syndrome (*scn1Lab^-/^*^-^) zebrafish, a model of genetic epilepsy. CQ (1 µM) significantly reduced locomotor activity (Fig. 3C), as well as the duration (Fig. 3D) and frequency (Fig. 3E) of epileptiform brain activity, in spontaneously seizing *scn1Lab^-/-^*zebrafish, compared with vehicle treatment (embryo medium, 1 % DMSO). As controls, *scn1Lab^+/+^* (wild-type) larvae or a mixture of wild-type and *scn1Lab^+/-^* (heterozygous) larvae, which do not exhibit seizures nor epileptiform activity when assessed, were used (Fig. 3C, D-E).

The recently introduced ASM perampanel, which inhibits glutamate receptor signaling (*28*) and hence also interferes with the glutamatergic system, exhibited comparable antiseizure activity (at 1 µM) in these zebrafish epilepsy models (*26*). Next, we assessed glutamate levels in zebrafish heads following CQ treatment of wild-type, EKP-treated- and spontaneous seizing Dravet syndrome (*scn1Lab^-/-^*) zebrafish via LC-MS/MS (Fig. 3F-H). Glutamate levels were consistently and significantly reduced by CQ treatment of wild-type, EKP-treated and Dravet syndrome zebrafish as compared to VHC controls. These findings show that CQ can reduce glutamate, the most abundant excitatory neurotransmitter, in the brain, which likely contributes to its antiseizure activity.

### Clioquinol reduces seizure duration in a mouse 6-Hz psychomotor seizure model and increases anti-inflammatory markers in the SSSE mouse epilepsy model

The antiseizure activity of CQ was further evaluated in the mouse 6-Hz psychomotor seizure model. Focal seizures initiated in animals using 6 Hz (44 mA) corneal stimulation are generally considered pharmaco-resistant as they are difficult to treat with currently available ASMs (*29*, *30*). Our results show a significantly reduced seizure duration 60 min after i.p. administration of 10 mg/kg (but not 5 mg/kg) CQ compared to VHC-treated mice (Fig. 4A). The observed reduction in seizure duration by CQ treatment is comparable to that induced by fenfluramine (5 mg/kg and 20 mg/kg) treatment in the 6-Hz model (fig. S4). Fenfluramine was used as a benchmark because this prior anti-obesity drug is currently being used as a novel ASM against DRE, including Dravet Syndrome, Lennox Gastaut and Sunflower syndrome (*31*, *32*).

**Fig. 4.**
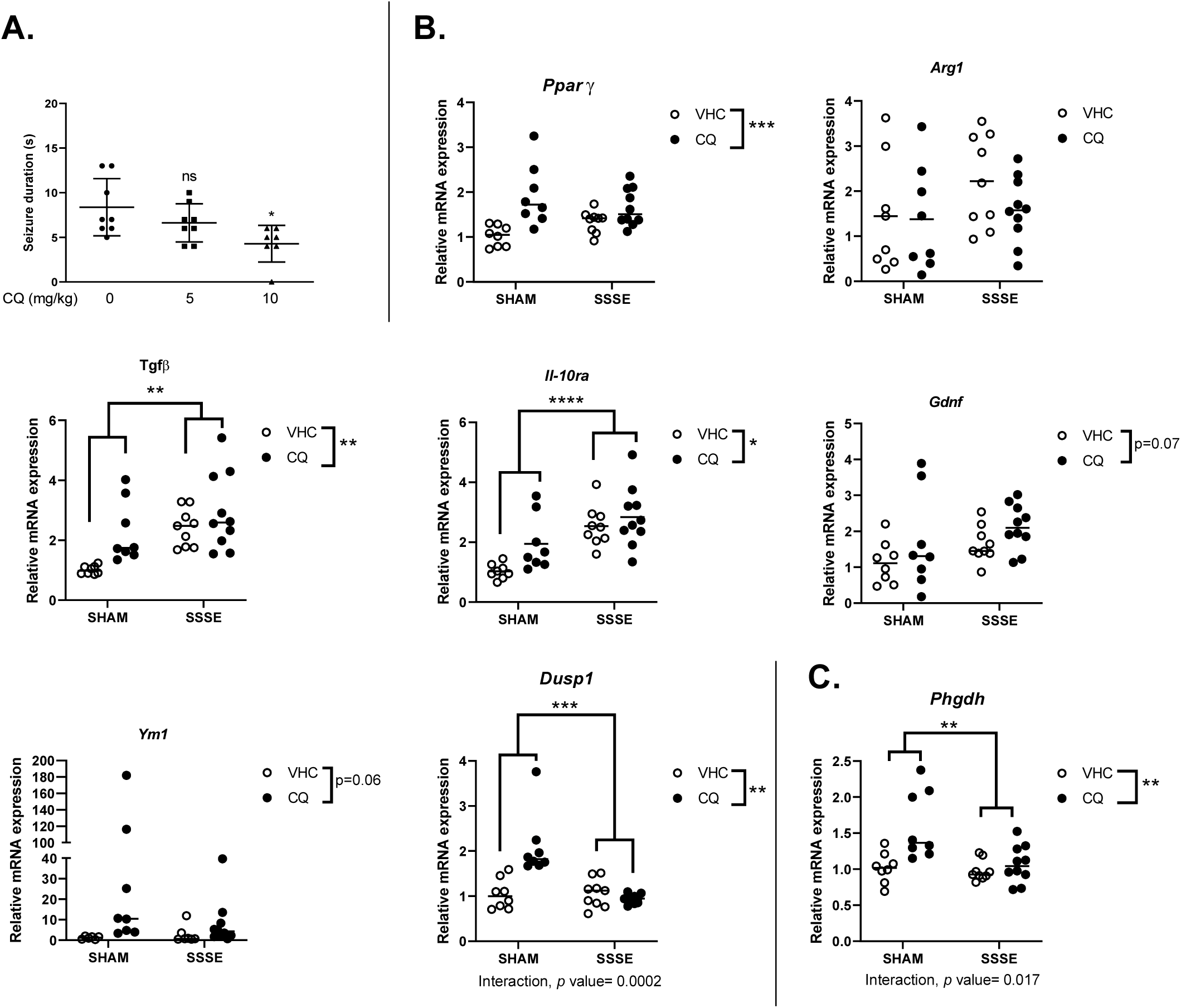
Antiseizure and anti-inflammatory activity analysis of clioquinol (CQ) in the mouse 6-Hz (44 mA) psychomotor and SSSE seizure models, respectively. (**A**) Drug-resistant psychomotor seizures were induced by electrical stimulation through the cornea, 60 min after i.p. injection of VHC (n = 8), CQ (5 mg/kg, n = 8) and CQ (10 mg/kg, n=7). Mean seizure durations (± SD) are depicted. Statistical differences: *p<0.05 by one-way ANOVA with Dunnett’s multiple comparisons test (**B, C**) The hippocampal mRNA expression of anti-inflammatory genes and Phgdh in CQ-treated SHAM and SSSE animals is represented as dot plots with median represented by a line. Statistical differences: ****p<0.0001, ***p<0.001, **p<0.01 and *p<0.05 by two-way ANOVA with Šídák’s multiple comparison test.

As PHGDH has been identified as a critical enzyme in driving macrophage polarization towards an anti-inflammatory state (*12*), we additionally investigated anti-inflammatory activity of CQ in a mouse model of Status Epilepticus (SE) (*33*). We assessed the expression of various anti-inflammatory markers 1 week after induction of SE via electrical stimulation, comparing CQ treatment (10 mg/kg/day) to vehicle. At the end of the experiment, unilateral hippocampi were collected for gene expression analyses (Fig. 4B,C). CQ-treated animals displayed a significant increase in the expression of *Phgdh,* as well as several anti-inflammatory cytokines and neuroprotective genes such as *Tfgb, Il-10ra*, *Dusp1*, *PPAR gamma*, *Gdnf* (p=0.07) and *Ym1* (p=0.06) when compared to VHC-treated mice. TGFb and IL10-ra were also upregulated by SE compared to sham SE, while the level of DUSP1, which is involved in the resolution of inflammation, was significantly reduced by SE. Several pro-inflammatory cytokines were significantly upregulated by SE, but no changes were induced by the CQ treatment except for a mild increase in the levels of *Il6* (p=0.06) and *Trem2* (p=0.06) (table S1). These data show that CQ may alter levels of anti-inflammatory mediators that could mediate immunomodulatory processes with the potential to promote repair mechanisms. It is currently not clear (i) how activation of PHGHD by CQ can result in increased expression of the Phgdh gene itself, and (ii) whether the observed CQ effects *in vivo* are resulting from PHGDH activation, increased expression of Phgdh or from a combination of both. In conclusion, by activating PHGDH, we target drug-resistant seizures as well as co-occurring neuroinflammation.

### Clioquinol reduces seizures in adolescents suffering from DRE

To test the safety and potential efficacy of CQ in DRE patients, we designed an open pilot signal finding study using CQ as an add-on therapy. The study also included a follow-up of pharmacokinetic parameters as well as the registration of potential adverse effects that might be linked to the development of subacute myelo-optic neuropathy (SMON; see discussion below) (*34*). Based on historical CQ pharmacokinetic studies in rodents (*35*, *36*) we calculated that the minimal effective CQ dose of 10 mg/kg i.p. in mice matches a 4 mg/kg CQ p.o dose in humans, which is much lower than doses associated with neurotoxicity/SMON (20-30 mg/kg/day) (*37*) and 4-fold lower than the CQ dose used for treatment of children with amoeba infections (15 mg/kg/day) (*38*).

In this study, three adolescents (CLIO-002, CLIO-003 and CLIO-004; Table 1) with severe DRE that failed at least 3 ASMs were included. All 3 subjects fulfilled the criteria of ‘Lennox Gastaut syndrome’. During the 4-week baseline period, a minimum of 4 countable convulsive seizures was a requirement for inclusion. After a 2-week low CQ dose exposure (1 mg/kg/day), patients received the effective CQ dose (4 mg/kg/day) for 6 weeks. The primary outcome was the seizure frequency decrease during the higher CQ dose exposure compared to baseline. Secondary outcomes included seizure severity based on National Hospital seizure Severity (NHS3) scale (*39*) and Quality of Life using the Personal Impact of Epilepsy Scale (PIES) (*40*) as well as safety.

**Table 1.**
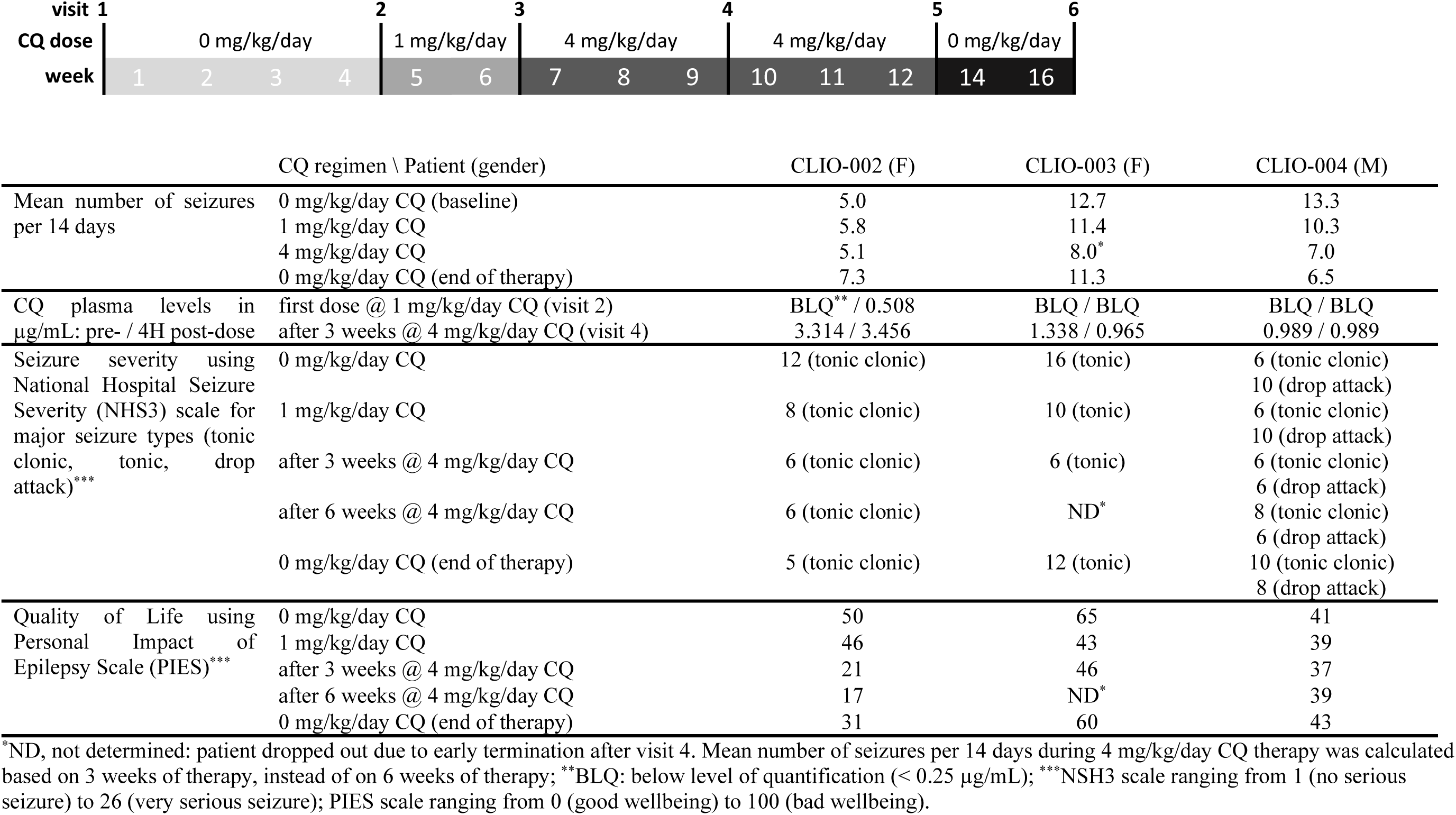
Trial design and primary and secondary outcome parameters of the clioquinol (CQ) proof-of-concept clinical study.

In two out of the three patients, a decrease of the seizure frequency upon receiving the effective CQ dose relative to baseline was seen (37,0% and 47,4% in patient 3 and 4 respectively). In view of the very refractory nature of their epilepsy, these results are promising and indicate a potential efficacy of CQ as add-on treatment in refractory epilepsy. No adverse events, including ophthalmological or cognitive, related to the treatment were reported during the trial. The seizure severity measured using NHS3 also decreased in all three patients. A positive impact of the new treatment on comorbidities and Quality of Life using PIES was seen in all three patients, and was most pronounced in patient CLIO-002, although this patient had no formal decrease in seizure frequency. Our data showed very low serum concentrations at the low CQ dose exposure (≤0.5 µg/mL) and stable levels at the higher dose (1-3.5 µg/mL), but still low compared to the CQ plasma level upon administration of a dose of 10 mg/kg/day in human (7.6 µg/mL) (*41*).

Repurposing CQ as an add-on ASM appears to be a valid option since this haloquinoline drug has been used for many years to treat traveler’s diarrhea, fungal and protozoal gastrointestinal tract infections (*14*). We acknowledge that a larger double-blind placebo-controlled trial will provide a definite answer on the antiseizure efficacy of CQ. Nevertheless, the three adolescents included in our signal finding study suffered from very refractory epilepsy, and a decrease in seizure frequency and severity is clinically meaningful. This was also reflected in the secondary outcomes: a positive impact on Quality of Life and less severe seizures were seen in all three patients. The efficacy might also have been underestimated as our data show that CQ plasma levels were low (1-3.5 µg/mL) in all patients.

Note that CQ overuse was considered a possible cause of SMON, which led to the drug’s withdrawal from the market (*34*, *42*). Specific single nucleotide polymorphisms (SNPs) in the antioxidant protein NQO1, mainly found in the Japanese population, have been proposed as a risk factor for developing SMON (*43*). Hence, we excluded the Asian population from the study. CQ induces oxidative stress specific to the retina, which can be counteracted by NQO1’s antioxidant activity. The most frequent human NQO1 polymorphism is NQO1*2 (C609T or Pro187Ser), which reduces NQO1 protein stability and cellular NQO1 activity. The striking connection of this polymorphism to the Japanese SMON cases is the much higher prevalence of this inactivating C609T NQO1 polymorphism in Japanese and Asian populations (>70-80%) compared to African or European populations (1-2%) and could be used for patient stratification in the future.

## Data Availability

All data produced in the present study are available upon reasonable request to the authors

## Acknowledgements

The authors wish to thank Jan Maes, Michèle Partoens and Jing Li for technical help during mouse and zebrafish experiments; Prof Wim M. De Borggraeve and Gert Steurs (Molecular Design and Synthesis, KU Leuven) for EKP synthesis; ‘scilled – scientific illustrations & design’ for designing Fig 1A; and Prof Patrik Verstreken (VIB-KU Leuven); Prof Pierre Vanderhaeghen (VIB-KU Leuven and Université Libre de Bruxelles) and Prof Patrick Kwan (University of Monash) for critical reading of the manuscript. We thank Dr Shauni Loopmans and Prof Dr Bart Ghesquière from the VIB Metabolomics Core Facility Leuven for the amino acid analysis.

## Funding

KU Leuven, IOFm/05/022 (KT) KU Leuven, C3/22/016 (KT) KU Leuven, IOFm/16/003 (AN)

KU Leuven, C14/17/107; C14/22/132 (LVDB)

National Health and Medical Research Council of Australia APP1141347 (BDS)

US Department of Defense W81XWH2010848 (BDS) and W81XWH-21-1-0927 (IA)

Fonds Wetenschappelijk Onderzoek (FWO), 1174523N (CG) and 11PGF24N (BF)

## Author contributions

Conceptualization: KT

Methodology: JT, KT, DC, AN, LL, PDW, JS, AV, PT, NJ, IA, CG, SP, VM

Investigation: JT, JS, AN, KL, EG, KK, KL, PT, IA, NJ, THT, MM, BDS, CG, BF, SP, VM

Visualization: KL, AN, KL, KK

Funding acquisition: KT, AN, IA, LVDB, BDS, CG, BF

Project administration: KT

Supervision: KT, AN, DC, LL, BPAC, PDW, AV, NJ, IA, TOB, LVDB, IE, CLL

Writing – original draft: KT

Writing – review & editing: all co-authors

## Data and materials availability

All data are available in the main text or the supplementary materials.

## Supplementary Materials

### Compound preparations

Clioquinol, broxyquinoline and CBR-5884 were purchased from Sigma-Aldrich (St Louis, USA) and chloroxine from TCI Europe (Zwijndrecht, Belgium); compound stock solutions were prepared in dimethyl sulfoxide (DMSO; VWR International, Belgium). Zinc sulphate heptahydrate, copper (II) sulphate pentahydrate and L-serine were from Sigma-Aldrich; stock solutions/compounds were prepared in milliQ water in 0.3X Danieau’s solution (1.5 mM HEPES, pH 7.6, 17.4 mM NaCl, 0.21 mM KCl, 0.12 mM MgSO_4_, and 0.18 mM Ca(NO_3_)_2_). Fenfluramine ((+-)FA) was a gift from prof. Berten Ceulemans (Child Neurology, University Hospital Antwerp, Belgium). Ethyl ketopentanoate (EKP) was synthesized by Molecular Design and Synthesis (Prof. Wim De Borggraeve, KU Leuven, Belgium) as described in (*26*).

### PHGDH enzymatic activity assay based on colorimetric NADH-based readout

To investigate PHGDH enzymatic activity upon drug treatment, human PHGDH (BPS bioscience, San Diego, USA) was used in combination with a specific colorimetric PHGDH activity assay kit (Abcam, Cambridge, UK) as described before (*44*). Prior to the assay, compound stock solutions were diluted in PHGDH assay buffer to obtain a 5% DMSO solution, and human PHGDH enzyme was diluted in assay buffer at a concentration of 0.38 mg/mL. DMSO background concentration was 0.5%. Absorbance at 450 nm (OD 450 nm) over time was measured as a readout for the amount of NADH generated through PHGDH activity. GraphPad Prism (version 6) software was used to calculate the area under the curve (OD 450 nm plotted against time (min)), representing PHGDH activity during 1h, and to perform statistical analysis. One-way ANOVA with Dunnett’s multiple comparisons test was performed to assign significant differences compared to the control condition (0.5% DMSO).

### PHGDH enzymatic assay in cellular context based on direct NADH readout

Recombinant human PHGDH and PSAT1 enzymes fused to N-terminal His_6_ tags were expressed in *E. coli* BL21(DE3) cells and purified to near homogeneity as previously described (*45*), except for an additional gel filtration step after the affinity purification. Gel filtration was carried out on an ÄKTA protein purifier system (ÄKTA Pure 25 M, Cytiva) using a Superdex 200 10/30 GL column (GE Healthcare) and a flow rate of 0.75 ml/min. The mobile phase contained 25 mM Tris, pH 7.5, and 150 mM NaCl. The protein concentration in the final purified preparation was determined by measuring A_280_ and using an extinction coefficient estimated with the ProtParam tool (RRID: SCR_018087). The protein purity for both purified enzymes was estimated at > 95% based on SDS-polyacrylamide gel electrophoretic analysis. The purified proteins were stored at −80°C after addition of 10% glycerol.

### Cell culture and preparation of deproteinized extracts

HAP1 cells (Horizon Discovery Group, Austria) were cultivated in Iscove’s Modified Dulbecco’s Medium (IMDM) supplemented with 100 units/ml of penicillin, 100 µg/ml of streptomycin, and 10% FBS (all medium reagents were from Gibco, Thermo Fisher Scientific) at 37°C and 5% CO_2_. The cells were passaged at least twice after thawing, before using them for experiments. Fourty-eight hours after seeding (1.5 x 10^6^ cells in 10 cm dishes containing 10 ml IMDM), cells were gently washed with 10 ml of pre-warmed (37°C) PBS and then scraped into 400 µl of pre-cooled (4°C) lysis buffer (50 mM HEPES, pH 7.1). Cell lysates were submitted to 3 freeze-thaw cycles, centrifuged at 17,000 x *g* for 30 minutes at 4°C, and the supernatants heated for 5 min at 95°C.

Denatured proteins were eliminated by another centrifugation at 17,000 x *g* for 30 minutes and 4°C and the final supernatants (referred to as “deproteinized cell extracts”) were used on the same day in activity measurements of recombinant hPHGDH.

### Enzymatic assay

The dehydrogenase activity of recombinant hPHGDH was measured spectrophotometrically (Multimode microplate reader Infinite 200 PRO M-FLEX, Tecan) at 37°C by monitoring the absorbance at 340 nm (formation of NADH; an extinction coefficient of ε = 6220 M^-1^ cm^-1^ was used for calculating initial velocities). We first incubated a mixture (total final volume 200 µl) containing 25 mM Tris (pH 9), 1 mM dithiothreitol, 1 mM MgCl_2_, 5 µM pyridoxal phosphate, 1 mM L-glutamate, 400 mM KCl, 0.5 mM NAD^+^, 100 µg/ml PSAT1, 10 µg/ml hPHGDH, and 100 µl of deproteinized HAP1 cell extract at 37°C for 10 minutes. The mixture also contained 20 µM or 200 µM Clioquinol (or 2% DMSO for the ‘0 µM Clioquinol’ condition). The reaction was started by addition of 300 µM 3-phosphoglycerate to determine initial velocities. Data analysis and plotting was performed with GraphPad Prism software (version 10.0.0). One-way ANOVA with Dunnett’s multiple comparisons test was performed to assign significant differences compared to the control condition.

### Protein expression and purification of sPHGDH

*E. coli* BL21(DE3) cells were transformed with sPHGDH (N terminal domains of PHGDH) encoded in pET28 (*19*). Colonies were transferred to LB medium containing kanamycin (50 µg/mL), and grown at 37°C. Protein expression was induced at OD by adding IPTG to a final concentration 0.5 mM, and growth was continued for 16 h at 20°C. The cells were harvested, and suspended in 100 mM Tris-HCl, pH 8.0, 150 mM NaCl, 1 mM DTT, 0.1 mM PMSF and it was sonicated on ice. The lysate was centrifuged, and then the supernatant was loaded onto 5 mL nickel-sepharose column (Qiagen). The nickel resin was washed with 50 mM Tris-HCl, pH 8.0, 150 mM NaCl, and after washing, eluted with 50 mM Tris-HCl, pH 8.0, 150 mM NaCl, 500 mM Imidazole. The elution fractions including sPHGDH were collected and it was concentrated to 3 mL using Amicon centrifugal filter units (Millipore). The concentrated protein was loaded onto a size-exclusion column (GE Healthcare) with 50 mM HEPES, pH 7.5, 100 mM NaCl, 1 mM EDTA.

### sPHGDH enzyme kinetics assessment

Assays were performed in 96-well transparent film bottom plates (Greiner) at 37°C. The reaction solution was 50 mM HEPES, pH 7.5, 100 mM NaCl, 1 mM EDTA, 1 mM NAD^+^, 0.5-5 mM 3-PG, 0.1 mM Resazurin, 7.5% DMSO, 50 µM sPHGDH, 0-50 µM Clioquinol, and the reaction volume 200 µl. CQ was not soluble under reaction conditions at higher concentrations. The enzymatic reaction was measured by detecting Resorufin produced by reduction of resazurin by produced NADH. The fluorescence of Resorufin (Ex: 550 nm / Ex: 580 nm) was measured by the plate reader (TECAN Safire). Measurements made with different enzyme samples were normalized by no CQ measurements. The initial rate was measured in three independent experiments and calculated by averaging. V_max_ values were calculated by Michaelis-Menten curve fitting using R.

### Determination of serine, glycine and glutamate levels in 4T1 breast cancer cell line and glucose tracing experiments

The tumor-derived 4T1 breast cancer cell line was kindly provided by the Fendt Lab (VIB-KU Leuven). 4T1 cells were thawed and plated in 10cm dishes (Sarstedt Inc 83.3902.300) in RPMI 1640 (Gibco™ 11875093) with 10% FBS (Biowest S1400-500), 1% Sodium Pyruvate 100mM (Gibco™ 11360070), 1% HEPES 1M (Gibco™ 15630080), 0,1% 2-mercaptoethanol (Sigma M3148) medium (Complete RPMI medium). RPMI 1640 medium deprived in glucose, L-glycine, L-glutamine and L-serine (Cell Culture Technologies LLC 1270RPM-0324) was prepared according to the manufacturer protocol using sodium hydrogen carbonate (Sigma S5761). To this media, ^12^C glucose (2000mg/L) (Sigma G7021), ^13^C_6_ glucose (2000mg/L) (Sigma 389374), L-glutamine (300mg/L) (Sigma G8540), 1% sodium pyruvate 100mM, 1% HEPES 1M, 0,1% 2-mercaptoethanol and 10% dFBS, generated by filtration of the previously used FBS using Slide-A-Lyzer (Thermo Scientific™ 66130, 66493), were added, generating serine and glycine deprived RPMI1640 culture medium (Special RPMI 1640 medium). 45.000 4T1 cells/well were plated in 6-well plates (CELLSTAR® 657160) in 2 mL/well of the complete RPMI 1640 medium. Cells were counted using the Countess™ 3 Automated Cell Counter (Invitrogen™ AMQAX2000). The day after plating the 4T1 cells in 6-well plates, the media was aspirated and, after a wash in DPBS, the media was replaced with the special RPMI 1640 depleted in serine and glycine. 3,055mg of Clioquinol (Sigma 24880) were dissolved in DMSO (Sigma D4540) to make a 10mM Clioquinol solution and added to the media at a final concentration of 5 μM and 7.5 μM. DMSO only was used as a control condition. After 24 h, the media was aspirated, the wells were thoroughly washed with DPBS and the 6-well plates were quenched by a 30 seconds contact with liquid nitrogen.

Metabolite extraction was performed using the methanol/chloroform method. The methanol extraction solution was prepared with 60% methanol (Fisher Scientific) in LC/MS grade water (Fisher Scientific) containing 6.67 μg/ml glutaric acid (Merck Sigma) and 6.67 μg/ml norvaline (Merck Sigma) as internal standard. Cells were resuspended in 800μL of −20° C-cold extraction solution and then 500μL of chloroform (Acros Organics NV) were added on top. Samples were vortexed for 10 min at 4° C and then centrifuged at 13,000 rpm at 4° C for 10 min. The upper phase containing the extracellular metabolites was transferred into fresh tubes and dried down overnight in a vacuum concentrator (Labconco) at 4°C. Dried metabolites were stored at −80° C. Protein interphase was also dried down and dissolved in 200μL 0.2M sodium hydroxide, then protein concentration was measured with the Pierce BCA Protein Assay Kit (Thermo Fisher Scientific 23225). Metabolites were measured using gas chromatography-mass spectrometry as previously described (*46*).

### Determination of glutamate and glycine levels in iPSC-derived astrocytes

#### iPSC line and culture

Control iPSCs were generated previously by genetic correction of an iPSC line derived from a 17-year-old male amyotrophic lateral sclerosis (ALS) patient using CRISPR-Cas9 (*47*). iPSCs were cultured on Geltrex (Gibco, A1413301) in Complete Essential 8 medium (Gibco, A1517001) with 1% penicillin/streptomycin (Gibco, 15070063).

#### Generation of iPSC-derived astrocytes

iPSCs were differentiated in mature astrocytes using the previously described protocol (*48*). In short, iPSCs were dissociated with collagenase type IV (Gibco, 10780004) to form embryoid bodies (EBs) in Corning ultra-low attachment flasks (Sigma-Aldrich, 734–4140) and kept in neuronal induction medium (NIM) consisting of Complete Essential medium with 1% penicillin/streptomycin supplemented with 0.1 μM LDN-193189 (Stemgent, 04–0074-02) and 10 μM SB431542 (Tocris, 1614) for the first week to support neuronal induction. On day 7, the EBs were plated on Geltrex-coated plates in neuronal maturation medium (NMM) containing 50% DMEM/F12 (Gibco, 11330032) and 50% Neurobasal medium (Gibco, 21103049) with 1% L-glutamine (Thermo Scientific, 25030–024), 1% penicillin/streptomycin, 1% N-2 supplement (Gibco, 17502–048) and 2% B-27™ without vitamin A (Gibco, 12587–010) supplemented with 10 ng/ml recombinant murine fibroblast growth factor (FGF)-2 (PeproTech, 450–33), 10 ng/ml recombinant human epidermal growth factor (EGF) (ProSpec, CYT-217), 0.1 μM LDN-193189 and 10 μM SB431542 to form neural rosettes and induce neural progenitor cell (NPC) expansion. NMM was changed every other day and NPCs were passaged using accutase (Sigma-Aldrich, A6964) when 100% confluency was reached. On day 16, neuronal NMM was switched to astrocyte differentiation medium (ADM) consisting of 90% Neurobasal medium, 1% penicillin/streptomycin, 1% N-2 supplement, 1% non-essentialamino acids (Gibcro, 11140050) and 0.8 μM ascorbic acid (Sigma-Aldrich, A4403) supplemented with 10 ng/ml FGF-2, 200 ng/ml recombinant human insulin like growth factor (IGF)-1 (Peprotech, 100–11), 10 ng/ml human Activin A (Gibco, PHC9564) and 10 ng/ml recombinant human Heregulinβ1 (Peprotech, 100–03) and changed every other day until day 25 to convert NPCs to astrocyte progenitor cells (APCs). On day 25 (d25/d+0), a glial switch occurs to commence the astrocyte maturation, which takes an additional 4 weeks. APCs were plated on Geltrex-coated plates for expansion in astrocyte maturation medium (AMM) consisting of 50% DMEM/F12, 50% Neurobasal medium, 1% non-essential amino acids, 1% N-2 supplement, 1% L-glutamine, 1% penicillin/streptomycin, 2% fetal bovine serum (FBS) (Gibco, 10270106), 0.8 μM ascorbic acid, and 1% sodium pyruvate (Gibco, 11360–070) supplemented with 200 ng/ml IGF-1, 10 ng/ml Activin A and 10 ng/ml Heregulinβ1 which was changed every other day. After 28 days (d+28) of maturation, mature astrocytes were plated in Geltrex-coated 12-well plates at 300.000 cells per well for glutamate and glycine determination.

#### Glutamate and Glycine determination

After 30 days of maturation (d+30), mature astrocytes were treated with 125 nM clioquinol, 62.5 nM clioquinol or 1% DMSO as vehicle for 50h in AMM without FBS containing 5 mM glucose (Sigma-Aldrich, G7021) and 2mM lactate (Sigma-Aldrich, 71718). After a wash with ice cold 0.9% NaCl solution, metabolites were extracted using 300 μL of an 80% methanol extraction buffer containing 2 μM of deuterated (d27) myristic acid as internal standard. Protein concentrations were determined using the micro BCA kit (23235, Pierce Biotechnology, Rockford, US). Following extraction, samples were centrifuged at 20.000×g for 20 min at 4 °C to remove precipitated proteins and insolubilities. Glutamate and glycine abundances were determined in the supernatant using mass spectrometry. In short, 10 µl of supernatant was loaded into a Dionex UltiMate 3000 LC System (Thermo Scientific Bremen, Germany) equipped with a C-18 column (Acquity UPLC-HSS T3 1. 8 µm; 2.1 x 150 mm, Waters) coupled to a Q Exactive Orbitrap mass spectrometer (Thermo Scientific) operating in negative ion mode. A step gradient was carried out using solvent A (10 mM TBA and 15 mM acetic acid) and solvent B (100% methanol). The gradient started with 5% of solvent B and 95% solvent A and remained at 5% B until 2 min post injection. A linear gradient to 37% B was carried out until 7 min and increased to 41% until 14 min. Between 14 and 26 minutes the gradient increased to 95% of B and remained at 95% B for 4 minutes. At 30 min the gradient returned to 5% B. The chromatography was stopped at 40 min. The flow was kept constant at 0.25 mL/min and the column was placed at 40°C throughout the analysis. The MS operated in full scan mode (m/z range: [70.0000-1050.0000]) using a spray voltage of 4.80 kV, capillary temperature of 300 °C, sheath gas at 40.0, auxiliary gas at 10.0. The AGC target was set at 3.0E+006 using a resolution of 140000, with a maximum IT fill time of 512 ms. Data collection was performed using the Xcalibur software (Thermo Scientific). The data analyses were performed by integrating the peak areas (El-Maven – Polly - Elucidata) and normalized to sample protein concentration. One-way ANOVA with Dunnett’s multiple comparisons test was performed to assign significant differences.

### Zebrafish EKP seizure model and Dravet syndrome epilepsy model

Adult zebrafish (*Danio rerio*) of AB strain (Zebrafish International Resource Center, Oregon, WA, USA) and *scn1Lab* strain (Dravet syndrome, a gift from Dr. Herwig Baier, Max Planck Institute of Neurobiology, Germany) were kept at 28°C, pH 6.5-7.8 and in a 14/10h light/dark regimen (standard aquaculture conditions). Upon natural spawning, fertilized eggs were selected and raised in embryo medium (0.3X Danieau’s solution) at 28°C in continuous light until 7 days post fertilization (dpf). The approval numbers for zebrafish experiments are 027/2019 (Ethics Committee of the University of Leuven) and LA1210261 (Belgian Federal Department of Public Health, Food Safety, and Environment). Compound stocks were diluted in embryo medium in a 96-well plate, containing one 7 dpf larva per well (resulting in 1% final DMSO background).

### Toxicity evaluation

The maximum tolerated concentration (MTC) of the compounds for the zebrafish larvae (in 1% DMSO) was determined as described previously (*49*). To determine MTC of CQ in combination with the Phgdh inhibitor CBR-5884, larvae were treated with a ½ MTC concentration of CQ and a concentration range of CBR-5884; MTC of the combination was determined according to the criteria in (*49*).

### Locomotor activity

To assess potential antiseizure activity of the compounds, AB and *scn1Lab* larvae were placed in the wells of a 96-well plate (1 larva/well) and treated with the appropriate compound in 100 µL embryo medium (1% DMSO) for 2h at 28°C in the dark. For the EKP assay, 100 µL vehicle (VHC; embryo medium, 1% DMSO) or EKP (600 µM in embryo medium, 1% DMSO) was added to the wells, resulting in a final EKP concentration of 300 µM before locomotion activity was assessed. For the Dravet syndrome assay, locomotion activity of *scn1Lab* larvae was assessed during a 40 min period where the first 30 min was considered as habituation. All locomotor activity was measured in an enclosed tracking device (ZebraBox Viewpoint, France). Locomotor activity in the EKP assay was expressed in “actinteg” values (ZebraLab software, Software Viewpoint, France), previously described as “the sum of all image pixel changes detected during the time window” (*26*). Data were plotted as movement (mean actinteg units/5 minutes relative to EKP-only treatment) during the 30 min recording interval. For the *scn1Lab^-/-^* zebrafish larvae, locomotor activity was expressed in “lardist” i.e total distance covered by the animal in large movements. Data was plotted as movement (mean lardist relative to the *scn1Lab^-/-^* larvae) during the last 10 min of a 40 min recording period. A one-way ANOVA with Dunnett’s multiple comparison test was performed to assign significant differences compared to reference condition.

### Electrophysiology

Noninvasive local field potential (LFP) recordings of the optic tectum (midbrain) of 7 dpf zebrafish larvae (*26*, *49*) were performed to measure epileptiform brain activities upon treatment with the compounds of interest (*e.g.* CQ, CBR-5884, a combination of CQ and CBR-5884, or L-serine). Larvae were treated with the compounds as described above for 2h. For the EKP assay, 100 µL VHC or 600 µM EKP (300 µM working concentration) was added, followed by 12 minutes of incubation in the dark before assessment. All larvae were immobilized in 2% low-melting-point agarose (Invitrogen) at room temperature and positioning of a single glass electrode containing artificial cerebrospinal fluid (ACSF; 124 mM NaCl, 2 mM KCl, 2 mM MgSO4, 2 mM CaCl2, 1.25 mM KH2PO4, 26 mM NaHCO3, and 10 mM glucose) above the optic tectum on the skin. Local field potential recordings of 10 minutes were performed as described. Data analysis was conducted with some adaptations. Briefly, Clampfit 10.2 software (Molecular Devices Corporation, USA and Matlab R2018 (MATrix LABoratory USA) software were used for visual inspection (for the Dravet syndrome assay) and execution of a power spectral density (PSD) analysis of the LFP recordings (for the EKP assay), respectively (*49*). PSD analysis was followed by normalization of the PSD estimates against the VHC control and the mean PSD per larva over a 10-90 Hz frequency range was calculated for each condition. A one-way ANOVA with Dunnett’s multiple comparison test was performed to assign significant differences between the treated groups of larvae and controls for each experiment. Significant differences in epileptiform brain activity between CBR-5884 treated larvae and VHC-treated control larvae were determined by means of an unpaired student t-test (two-tailed).

### Determination of glutamate levels in zebrafish heads

To determine the amount of glutamate in zebrafish heads, 7 dpf larvae (12/condition) were treated with or without 1 µM CQ in 1 mL of VHC (embryo medium, 1 % DMSO) for 2h at 28°C in the dark, followed by the addition of 1 mL VHC or 1 mL EKP solution (600 µM in embryo medium, 1% DMSO). After 1 min or 6 minutes respectively, larvae were washed in embryo medium, and heads were isolated using a scalpel under a dissecting microscope. Further sample preparation was performed by homogenizing 10 heads/condition in 120 µl of antioxidant medium (0.27 mM Na_2_EDTA.2H_2_O, 0,1 M acetic acid, 3.3 mM L-cystine, 12.5 µM ascorbic acid) by twisting and moving a microtube homogeniser up and down 50 times at 4°C. After centrifugation (12,000 g, 15 min, 4°C), 100 µl of the supernatant was kept at −80°C until analysis. Glutamate concentration in the samples was analyzed by LC-MS/MS. The glutamate levels in zebrafish head homogenates are shown as normalized glutamate amounts for the different treatments. Unpaired student t-tests were performed to assign significant differences in Fig 3. (panels F and G), while a two-way ANOVA with Šídák’s multiple comparisons test was performed to assign significant differences in Fig. 3 (panel H).

### Mouse 6-Hz psychomotor seizure model

Male Naval Medical Research Institute (NMRI) mice, provided by Charles River Laboratories, were housed (5 mice/cage) and maintained as described in (*49*) until the experiment was conducted. Mouse experiments were approved by the Ethics Committee of the University of Leuven (approval number 027/2017) and by the Belgian Federal Department of Public Health, Food Safety, and Environment (approval number LA1210261).

Antiseizure activities of CQ and fenfluramine were assessed in the mouse 6-Hz psychomotor seizure model as described previously (*49*). Briefly, NMRI mice were randomly divided into different treatment groups and 500 µL (adjusted to the individual weight) of vehicle (VHC; 0.5% sodiumcarboxymethylcellulose (NaCMC)/Tween80 in 0.9% NaCl for CQ experiments and DMSO/PEG200 (50:50) for fenfluramine experiments) or treatment (CQ or fenfluramine dissolved in VHC) was injected intraperitoneally (i.p.). 1h after injection, corneas were moisturized by an ocular anesthetic (lidocaine, 0.5%) and psychomotor seizures were induced by corneal electrical stimulation (6 Hz, 0.2 ms rectangular pulse width, 3s duration, 44 mA) using an ECT Unit 5780 (Ugo Basile, Comerio, Italy). Typical characteristics of psychomotor seizures were observed by experienced researchers. Initially observed seizure durations were confirmed or corrected upon blinded video analysis. Antiseizure activity is shown as seizure duration for the different CQ doses, represented as dot plot with median represented by a line. A one-way ANOVA with Dunnett’s multiple comparison test was performed to assign significant differences compared to the VHC control treatment.

### SSSE mouse epilepsy model

Seven-week-old, C57bl/6/j mice bred at the Alfred Medical Research & Education animal facility were used for the gene expression study. The experiments were approved by the local Animal Ethics Committee (E/2077/2021/M). Mice were surgically implanted with three extradural screw electrodes (a ground, reference and an active electrode) and one bipolar stimulating electrode (PlasticsOne, USA) into the right ventral hippocampus (coordinates: AP: −3.00; ML: –3.00; and DV: 2.80). Surgery was conducted under isoflurane anesthesia (5% for induction; and 1-2 % for maintenance) with provision of presurgical analgesia to alleviate pain (Carprofen 5 mg/kg & Buprenorphine 0.5mg/kg, s.c). The electrode assembly was held in place with dental acrylic (Vertex Dental, The Netherlands). An after-discharge threshold was established as previously described in (*50*). Subsequently, mice received electrical stimulation for 90 min duration, 100-ms trains of 1-ms alternating current pulses (50 Hz) at a suprathreshold current intensity. At the end of 90-min stimulation, mice were monitored for another 150 min following which SSSE was terminated with diazepam (*51*). Mice received either CQ (5 mg/kg twice daily i.p.) or vehicle (VHC) injections for one week. CQ was suspended in 5% dimethyl sulfoxide and 20% Kolliphor RH40 in 0.01M PBS.

At the completion of treatment, hippocampi were dissected and immediately frozen on dry ice and stored at –80°C. mRNA was extracted using a Nucleospin RNA Plus kit (Machery-Nagel) and cDNA synthesis was performed using the Omniscript RT Kit (QIAGEN). The real-time quantitative PCR was completed using high-throughput gene expression analysis based on microfluidic dynamic arrays (48.48 Dynamic array IFC) via the Medical Genomic Facility (MHTP) at the Monash Health Translational Precinct (Clayton, Australia). Taqman gene expression assays (Thermo Fisher Scientific, USA) were obtained as 20x forward and reverse primer and probe mixes (table S2). Each primer was at a concentration of 18 μM and probe at concentration of 4 μM. The final concentration of each assay was 0.2X (180 nM). Data were analyzed with Fluidigm real-time PCR analysis software (V4.1.1, South San Francisco, USA) using the 2^-ΔΔCT^ method, with gene expression levels for each sample normalized to the geometric mean of housekeeping genes including GAPDH, ACTB, HPRT 1 and PPIA. CT values of these housekeeping genes were not impacted by experimental condition. All mRNA expression levels were normalized to values of control animals. The relative mRNA expression levels of genes of interest in CQ-treated SHAM and SSSE animals are represented as dot plots with median represented by a line. A two-way ANOVA with Šídák’s multiple comparison test was performed to assign significant differences between the VHC versus CQ as well as SHAM versus SSSE animals.

### Clioquinol Clinical Study

#### Study population and Study Drug

Major inclusion criteria for participation in the open label study (registered as EUDRACT 2020-004511-27): informed consent; a diagnosis of DRE as defined by the International League against Epilepsy, age 12 - 18 years and a minimum of 4 convulsive seizures in a 4-week baseline period. CQ was formulated and manufactured by ACE Pharmaceuticals (Zeewolde, The Netherlands) as a 100 mg/mL syrup. The study was an add-on open label pilot Phase II trial.

#### Study Design

Primary readout was seizure frequency/2 weeks. Prior to the administration of CQ, seizure frequency/2 weeks was recorded for 2-4 weeks (baseline). CQ was administered orally BID, first at a low dose for 2 weeks (1 mg/kg/day) followed by a higher dose for 6 weeks (4 mg/kg/day). The primary efficacy variable was change in seizure frequency/2 weeks during high dose regimen relative to baseline. During the trial, the concomitant antiseizure medication (ASM) was kept stable (max 3 ASMs). Secondary outcome measures included change of seizure severity, impact on daily life and Quality of Life.

#### Study procedures

Study visits were at T0 start baseline, start low dose time 0, start high dose (time 0+2 weeks), safety visit (time 0+ 5 weeks), end of study (Time 0 + 8 weeks). At weeks 2, 4 and 7, CQ plasma levels were assayed by means of high-performance liquid chromatography at BioNotus, Belgium. The effect on seizure severity, overall impact of seizures, medication side effects, comorbidities, and overall QoL, using the standardized questionnaires; National Hospital Seizure Severity Scale (NHS3) (*39*) and Personal Impact of Epilepsy Scale (PIES) (*40*).

#### Safety measures

Standard adverse event reporting was conducted. Clinical exam, including visual screening (questionnaire and clinical exam) and motor examination were done at all visits.

### Statistical analysis

All statistical analyses were performed using GraphPad Prism 9.5.0 software, and data are presented as mean ± standard deviation (SD). Outliers were removed by means of the ROUT method (Q=1%). Results were considered to be statistically significant if the adjusted P value was p<0.05 (*); p≤0.01 (**); p≤0.001 (***) and p≤0.0001 (****).

---

**Fig S1.**
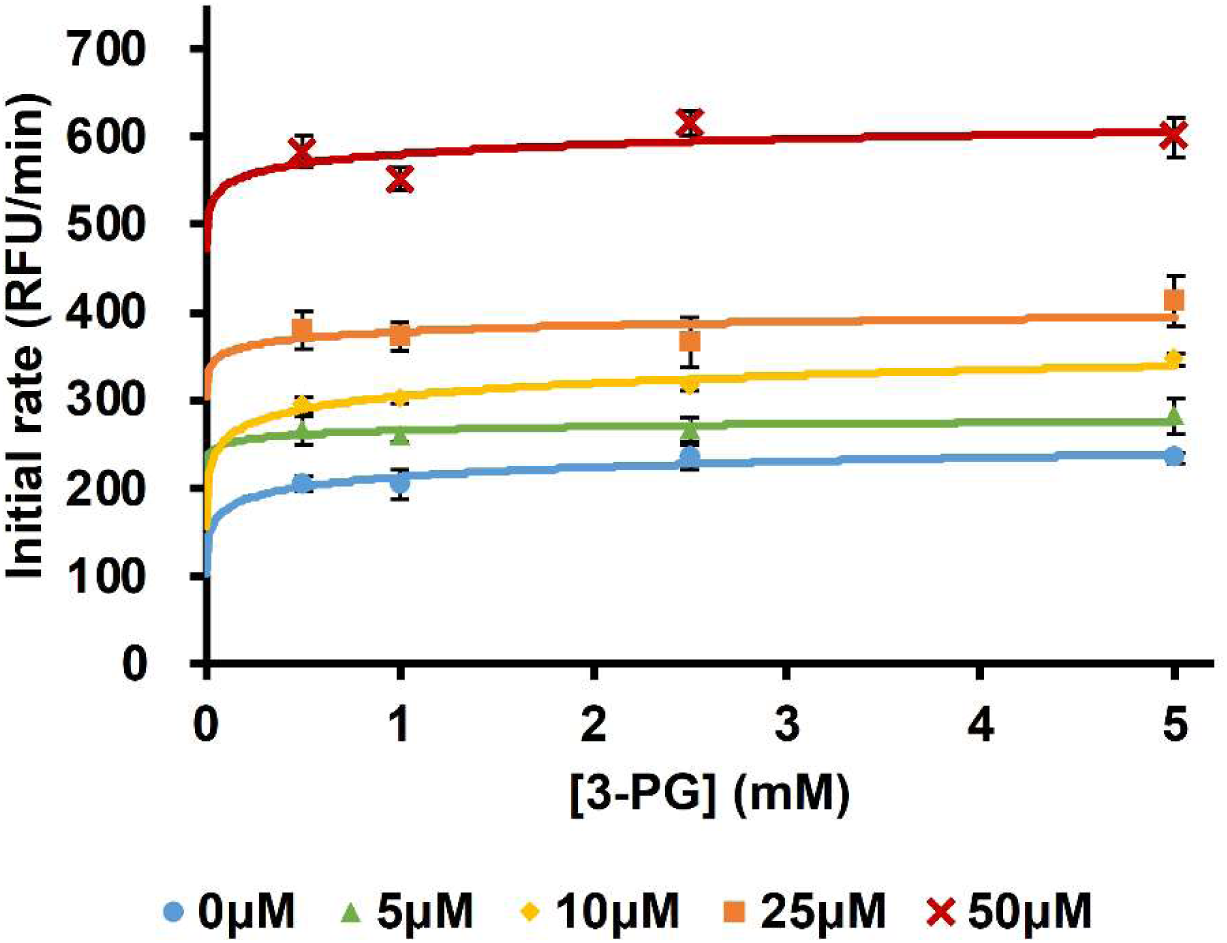
Concentration dependency of sPHGDH activation by CQ. The catalytic activity of sPHGDH in the presence of 0-5 mM 3-phosphoglycerate (3-PG) was investigated by following the NADH-induced colorimetric change of resazurin. The initial rate was calculated in the first 15 minutes at the linear range of the reaction in 3 independent experiments and is indicated as mean ±SD.

**Fig. S2.**
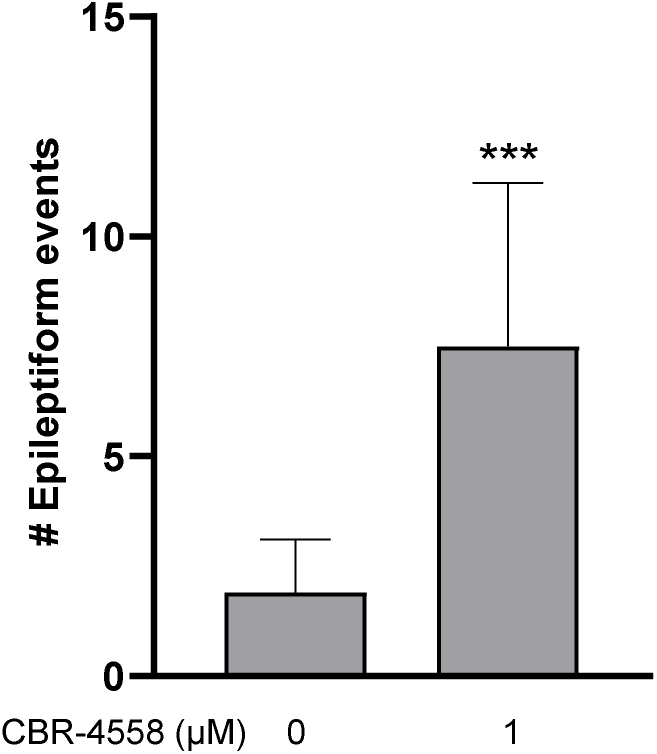
Effect of Phgdh inhibitor CBR-5884 on epileptiform brain activity in 7 dpf zebrafish larvae. Electrophysiological antiseizure activity (10 min non-invasive local field potential recording) was expressed in number of epileptiform events +/- SD; polyspiking events (≥ 3 spikes) with ≥ 3 times the amplitude of the baseline and lasting ≥ 50ms. Incubation time was 45 minutes. Number of recordings analyzed were VHC (n= 10), 1 µM CBR-5884 (n=10). Statistical differences: ***p<0.001 by unpaired Student’s t-test.

**Fig. S3.**
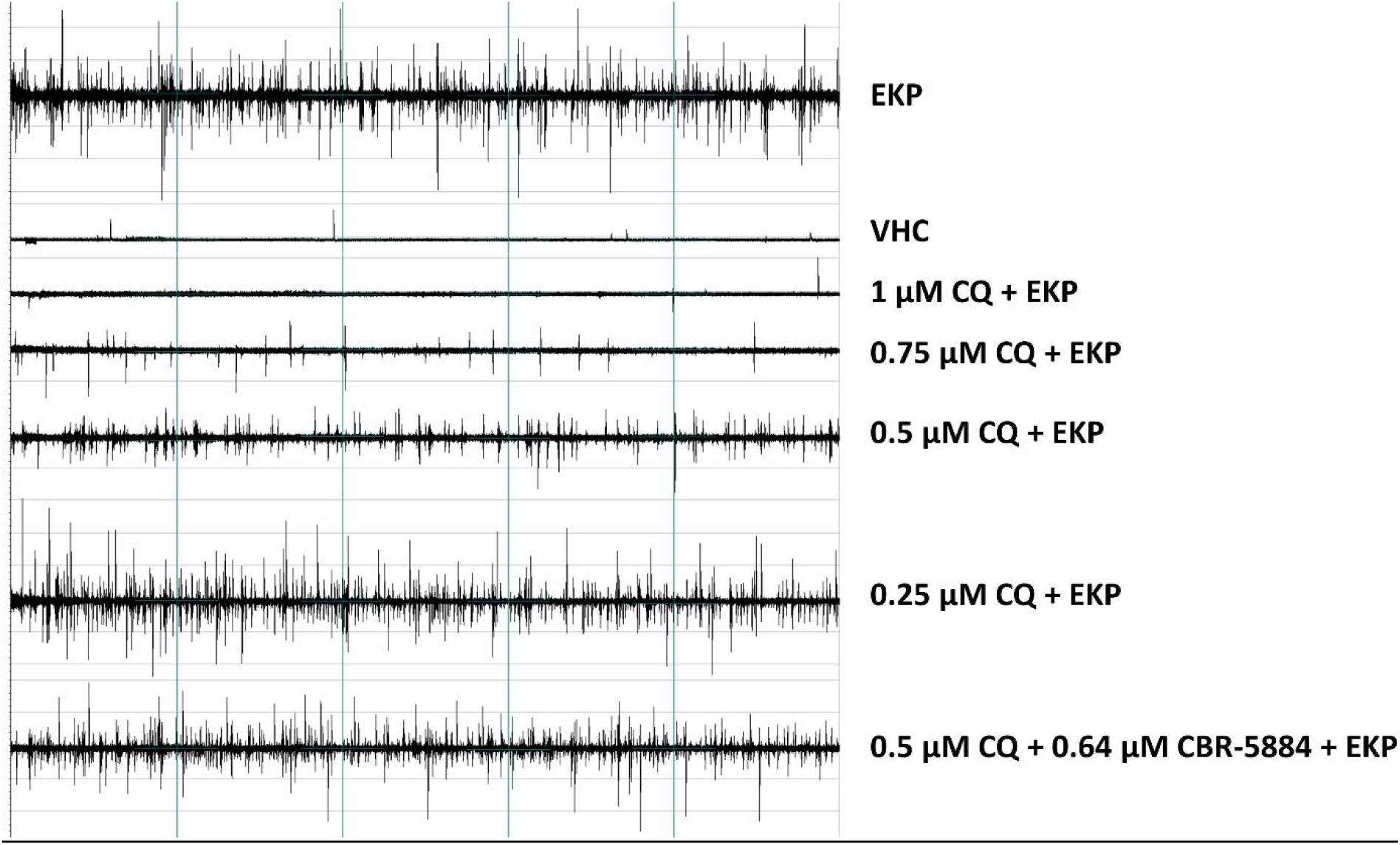
Representative LFP recordings of 7 dpf zebrafish. Zebrafish were treated with 1% DMSO (VHC) or 0.25 – 1 µM clioquinol, in the absence or presence of 0.64 µM Phgdh inhibitor CBR-5884, followed by the addition of EKP.

**Fig. S4.**
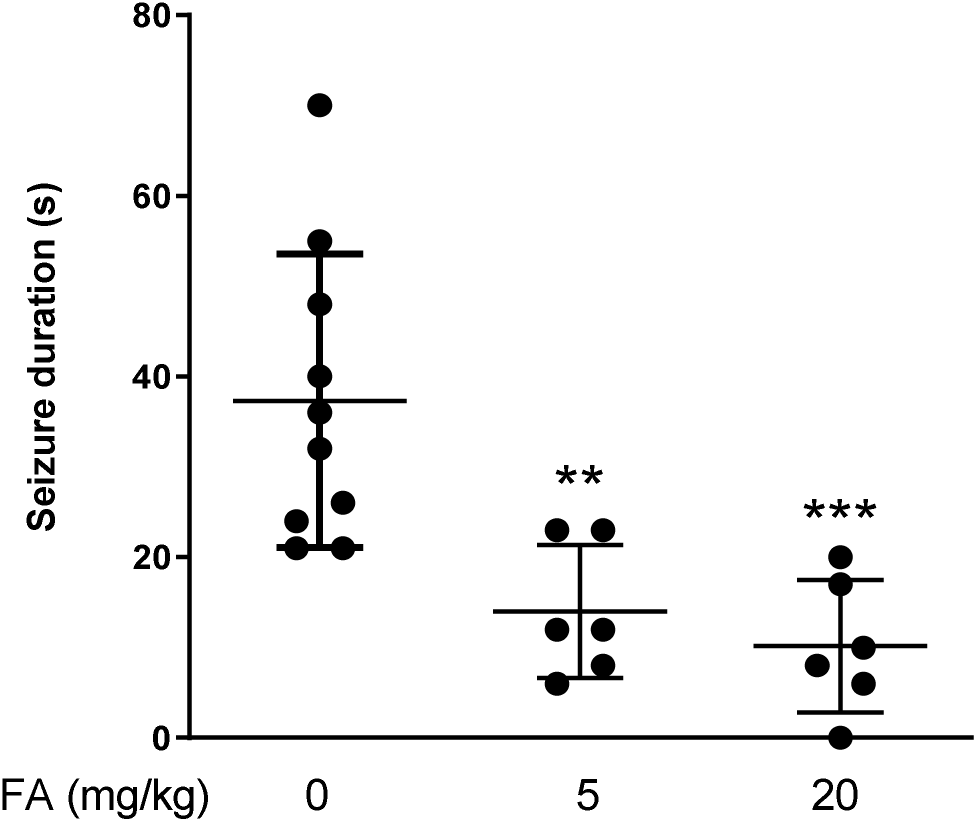
Antiseizure activity analysis of fenfluramine (FA) in the mouse 6-Hz (44 mA) psychomotor seizure model. Drug-resistant psychomotor seizures were induced by electrical stimulation through the cornea, 60 min after i.p. injection of vehicle (VHC, n = 9), FA (20 mg/kg, n = 6) and FA (20 mg/kg, n=6). Mean seizure durations (±SD) are depicted. Statistical differences: ***p<0.001 and **p<0.01 by one-way ANOVA with Dunnett’s multiple comparisons test.

**Table S1.**
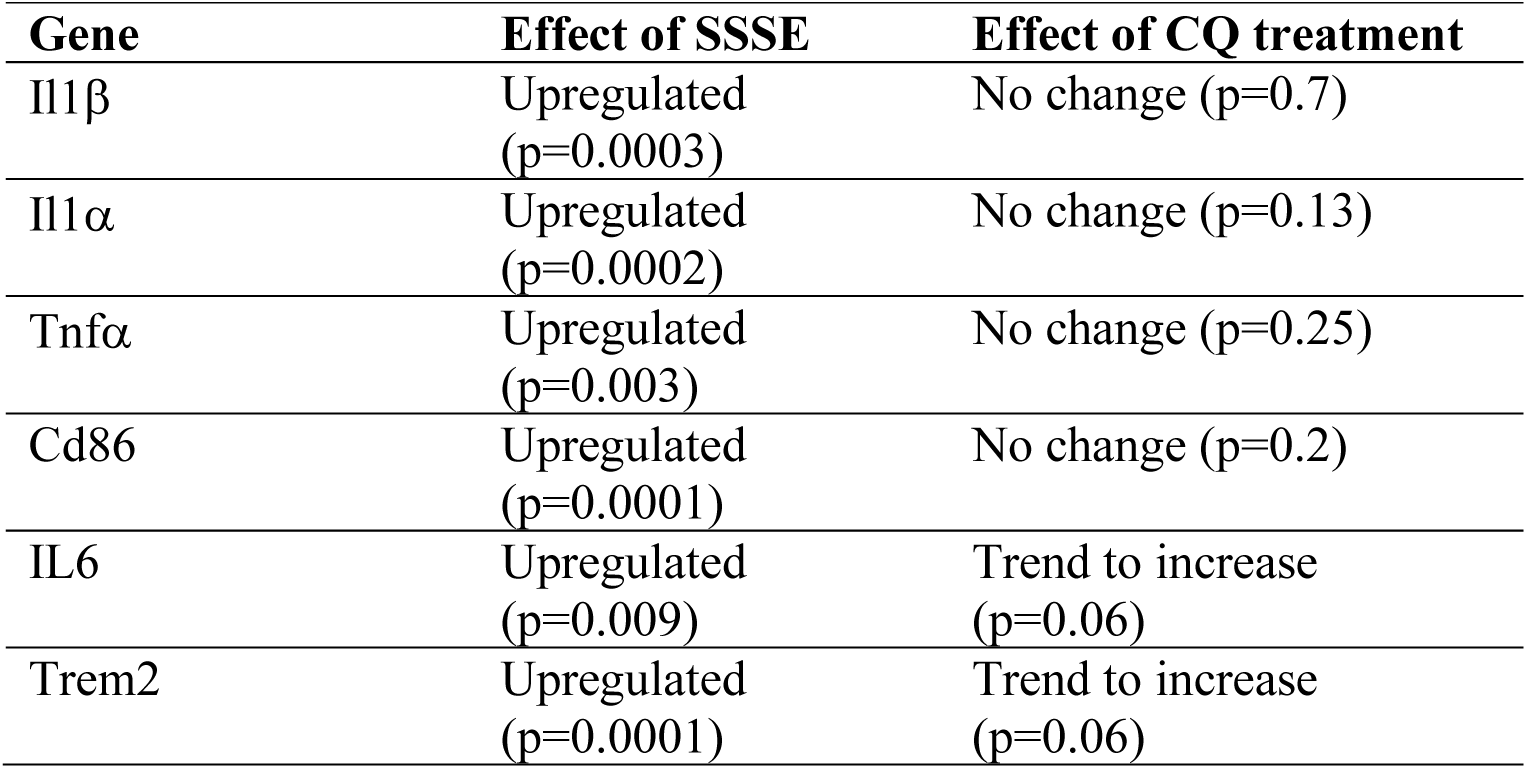
Effect of CQ on pro-inflammatory genes.

**Table S2.** Primers and corresponding Taqman assay ID.

| **Gene** | **Taqman assay ID** |
| --- | --- |
| *Phgdh* | Mm01623589_g1 |
| *Tgfb* | Mm01227699_m1 |
| *Arg1* | Mm00475988_m1 |
| *Il-10ra* | Mm00434151_m1 |
| *Gdnf* | Mm00599849_m1 |
| *PPARg* | Mm01184322_m1 |
| *Dusp1* | Mm00457274_g1 |
| *Ym1* | Mm00657889_mH |
| *Il1b* | Mm00434228_m1 |
| *Il1a* | Mm00439620_m1 |
| *Tnfa* | Mm99999068_m1 |
| *Cd86* | Mm00444543_m1 |
| *Il6* | Mm00446190_m1 |
| *Trem2* | Mm04209424_g1 |
| *Gapdh* | Mm99999915_g1 |
| *Actb* | Mm00607939_s1 |
| *Hprt1* | Mm00446968_m1 |
| *Ppia* | Mm02342430_g1 |

